# Proteomic Aging Clocks and the Risk of Mortality among Longer-Term Cancer Survivors in the Atherosclerosis Risk in Communities (ARIC) Study

**DOI:** 10.1101/2024.07.09.24309726

**Authors:** Shuo Wang, Zexi Rao, Anne H. Blaes, Josef Coresh, Corinne E. Joshu, James S. Pankow, Bharat Thyagarajan, Peter Ganz, Weihua Guan, Elizabeth A. Platz, Anna Prizment

**Affiliations:** Department of Laboratory Medicine and Pathology, University of Minnesota, Minneapolis, MN; Division of Biostatistics and Health Data Science, University of Minnesota, Minneapolis, MN; Division of Hematology, Oncology and Transplantation, University of Minnesota, Minneapolis, MN; Departments of Population Health and Medicine, NYU Grossman School of Medicine, New York, NY; Department of Epidemiology, Johns Hopkins Bloomberg School of Public Health, Baltimore, MD; Sidney Kimmel Comprehensive Cancer Center at Johns Hopkins, Baltimore, MD; Division of Epidemiology and Community Health, University of Minnesota, Minneapolis, MN; Division of Cardiology, Zuckerberg San Francisco General Hospital and Department of Medicine, University of California, San Francisco, CA

## Abstract

**Background:** We constructed a new proteomic aging clock (PAC) and computed the published Lehallier’s PAC to estimate biological age. We tested PACs’ associations with mortality in longer-term cancer survivors and cancer-free participants.

**Methods:** ARIC measured 4,712 proteins using SomaScan in plasma samples collected at multiple visits, including Visit 5 (2011-13), from 806 cancer survivors and 3,699 cancer-free participants (aged 66-90). In the training set (N=2,466 randomly selected cancer-free participants), we developed the new PAC using elastic net regression and computed Lehallier’s PAC. Age acceleration was calculated as residuals after regressing each PAC on chronological age after excluding the training set. We used multivariable-adjusted Cox proportional hazards regression to examine the associations of age acceleration with all-cause, cardiovascular disease (CVD), and cancer mortality.

**Results:** Both PACs were correlated with chronological age [r=0.70-0.75]. Age acceleration for these two PACs was similarly associated with all-cause mortality in cancer survivors [hazard ratios (HRs) per 1 SD=1.40-1.42, p<0.01]. The associations with all-cause mortality were similar in cancer survivors and cancer-free participants for both PACs [p-interactions=0.20-0.62]. There were also associations with all-cause mortality in breast cancer survivors for both PACs [HRs=1.54-1.72, p<0.01] and colorectal cancer survivors for the new PAC [HR=1.96, p=0.03]. Additionally, the new PAC was associated with cancer mortality in all cancer survivors. Finally, HRs=1.42-1.61 [p<0.01] for CVD mortality in cancer-free participants for two PACs but the association was insignificant in cancer survivors perhaps due to a limited number of outcomes.

**Conclusion:** PACs hold promise as potential biomarkers for premature mortality in cancer survivors.

## Introduction

The increased life expectancy in the US and improved cancer survival have significantly increased the number of cancer survivors, which is projected to reach 26 million by 2040.^1^ Cancer survivors experience accelerated aging; so their biological age is older than their chronological age and they experience premature death.^2–5^ Therefore, there is a need to develop biomarkers that can estimate biological age and predict mortality risk in cancer survivors.

To estimate biological age, researchers have developed aging clocks using DNA methylation profiles (epigenetic clocks), gene expression, circulating proteins, or other biomarkers.^6^ A few studies applied epigenetic clocks to study the aging process in cancer survivors;^7–10^ however, the underlying mechanisms of aging-related changes in DNA methylation sites remain unclear.^11^ Proteomic aging clocks (PACs), constructed using circulating proteins, are appealing because proteins are most proximal to diseases and thus may provide more information on age-related pathologies compared to other types of aging clocks.^11–13^ PACs were associated with mortality risk in the general population in our previous study^14^ and published studies.^15,16^ However, no studies have tested PACs and mortality in cancer survivors.

In this study, we examined the associations between PACs and mortality in long-term cancer survivors and cancer-free participants within the Atherosclerosis Risk in Communities (ARIC) study. ARIC has measured 4,712 proteins using SomaScan in plasma samples collected at multiple visits, including Visit 5 (2011-13) from participants aged 66-90 years. We developed a new PAC using proteins measured in cancer-free participants. We also computed the published PAC developed by Lehallier [2020], called here Lehallier’s PAC.^17^ We examined the associations of these PACs with all-cause mortality and cardiovascular disease (CVD) mortality in cancer survivors and cancer-free participants, and with cancer mortality in cancer survivors. Our hypothesis is that PACs are positively associated with the risk of mortality in cancer survivors and cancer-free participants.

## Methods

### Study population

The ARIC study (RRID: SCR_021769) is an ongoing cohort that enrolled 15,792 mostly White and Black females and males in 1987-89 from four study centers: Maryland, Minnesota, Mississippi, and North Carolina. Institutional review boards at each center approved the ARIC study, and all study participants provided written informed consent.^18,19^ By 2023, ten visits have been completed. Participants have undergone annual telephone follow-ups in 1987-2012 and semi-annual after 2012. Response rates were 83%-99% for follow-up calls among living participants who consented to be contacted (**Supplemental Methods**).^19^

### Ascertainment of cancer cases, death, and participants’ characteristics

Cancer cases were ascertained through 2015 using state cancer registries at the four study centers, complemented by the abstraction of medical records and hospital discharge summaries.^19^ For most common cancers, such as breast, colorectal, lung, and prostate cancers, stage at diagnosis was determined using the pathologic TNM stage (tumor extent, lymph node involvement, presence of metastasis) from cancer registries or medical records. In instances where pathologic TNM stage was not available, stage at diagnosis was determined according to Surveillance, Epidemiology, and End Results (SEER) summary stage (**Supplemental Methods**).^19^

Deaths were ascertained through linkage to National Death Index.^20^ Deaths due to CVD and cancer were defined based on underlying cause of death (**Supplemental Methods**). Participants’ characteristics of interest included chronological age, sex, race, study center, education, smoking status, alcohol intake, body mass index (BMI), aspirin use, diabetes, CVD, stage at diagnosis (for most common cancers), and estimated glomerular filtration rate (eGFR) (**Supplemental Methods**).

### Protein measurement

SomaScan measured 4,712 proteins in plasma samples collected at Visit 2 (1990-92), Visit 3 (1993-95), and Visit 5 (2011-13). The details of SomaScan and the data normalization process have been described previously.^21–23^ Bland-Altman coefficient of variation (CVBA) for split samples was 6% at Visit 2, 12% at Visit 3, and 7% at Visit 5 (**Supplemental Methods**). Protein levels, expressed in relative fluorescent units (RFU), were log2-transformed to correct for skewness.

### Statistical analysis

PACs were constructed using R (version 4.1.2, package “glmnet”). All other analyses were performed using SAS version 9.4 (SAS Institute Inc, Cary, NC). Statistical significance was considered if a two-sided p-value <0.05.

Among 5,183 White and Black participants with protein measures at Visit 5, we excluded participants with prevalent cancer at enrollment and those with a cancer diagnosis within two years before their blood collection at Visit 5 (N=678), in order to minimize the impact of active cancer treatment on protein levels. This resulted in 806 long-term cancer survivors and 3,699 cancer-free participants. PACs were constructed among cancer-free participants in the training set (two-thirds randomly selected cancer-free participants). The remaining cancer-free participants were used as the test set (**Figure 1**).

**Figure 1.**
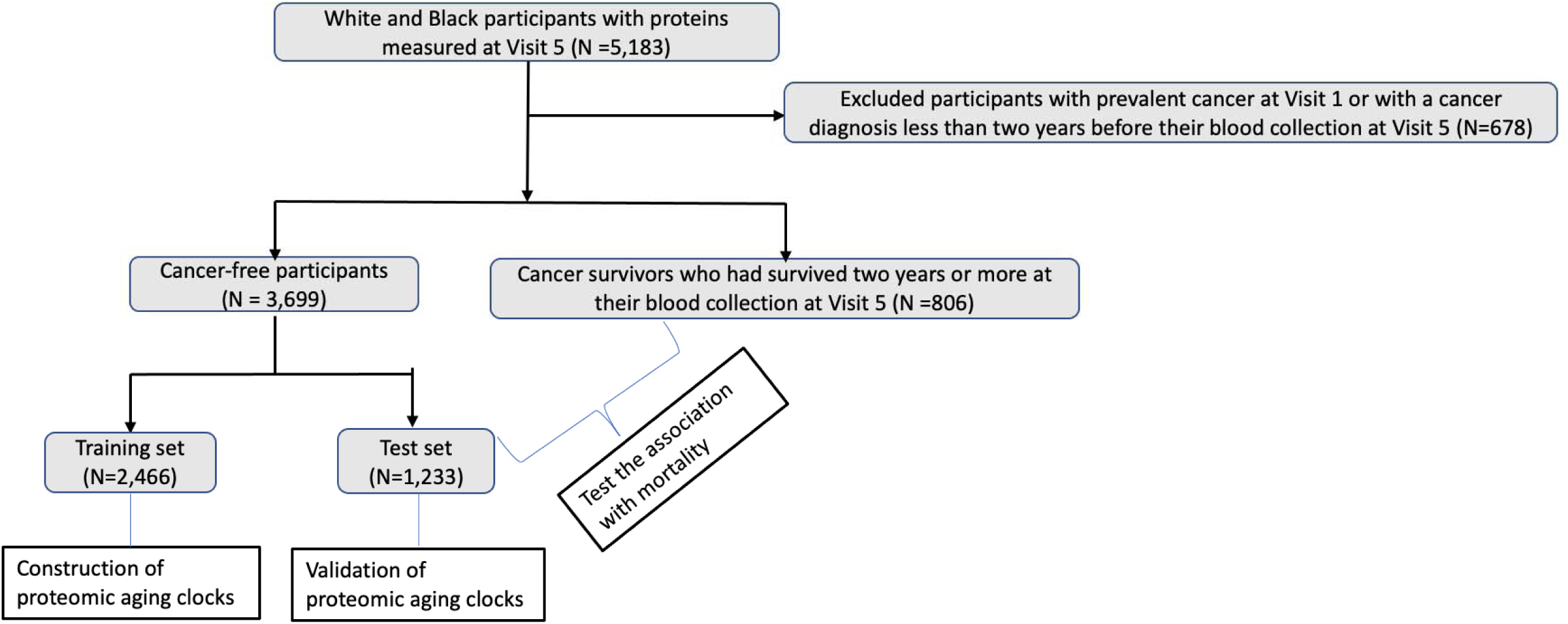
Study population.

Following the methodology used in previous studies,^16,24^ we applied elastic net regression to train the new PAC against chronological age in the training set (**Supplemental Methods**). Elastic net regression selected 619 aptamers for the new PAC: 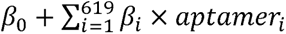, where *aptamer_i_* represents the level of *ith* aptamers, and estimated the intercept (β_0_) and non-zero weights (β_i_). To compute Lehallier’s PAC, we applied Ridge regression to estimate ARIC weights for the available aptamers in ARIC (**Supplemental Methods**). We did not use the published weights because ARIC included only 415 out of the 491 aptamers reported in Lehallier’s PAC.^17^ To internally validate these PACs, among cancer-free participants in the test set, we calculated Pearson correlation (r) between PAC and chronological age as well as median absolute error (MAE).

To capture the effects of PAC independent of chronological age, we calculated age acceleration for each PAC as residuals after regressing PAC on chronological age.^25^ We examined the distributions of participants’ characteristics at Visit 5 across tertiles of age acceleration in cancer survivors and cancer-free participants (test set). We used Cox proportional hazards regression to estimate hazard ratios (HR) and 95% confidence intervals (CIs) for all-cause mortality and CVD mortality in cancer survivors and cancer-free participants (test set), and cancer mortality in cancer survivors in relation to age acceleration (continuous). For CVD and cancer mortality, we treated deaths from other causes as competing events using the Fine and Gray method.^26,27^ Participants were followed from Visit 5 until death, censoring, or the end of follow up (December 31, 2017 for Mississippi participants or December 31, 2019 for participants from other centers), whichever occurred first. The proportional hazards assumption, assessed by modeling an interaction between age acceleration and follow-up time, met in all regression models. We adjusted for chronological age, sex, race-center, education, BMI, smoking status, alcohol intake, aspirin use, CVD, diabetes and eGFR (fully-adjusted model) (**Supplemental Methods**). We did not adjust for stage at diagnosis due to the incomplete information in all cancer survivors, but we adjusted for stage in the analysis of survivors of the most common cancers, namely breast, prostate, and colorectal cancers. We examined the association between age acceleration and all-cause mortality in survivors of these cancers in the fully adjusted model and an additional model that further adjusted for stage. Survivors of other cancers, including lung cancer (N<25), were not examined due to limited numbers.

We conducted three exploratory analyses. First, we used the KEGG database to identify pathways related to proteins in the new PAC. Second, we examined whether sex and race modified the association between age acceleration and all-cause mortality. Due to small numbers of CVD and cancer deaths, we did not stratify by sex and race in the associations with these outcomes. Third, we examined whether cancer status modified the association between age acceleration and all-cause mortality using a time-dependent model. To conduct this analysis, we used proteins measured at Visit 2 (1990-92) and Visit 3 (1993-95) and developed two additional PACs. In the time-dependent model, we followed participants from Visit 2, updated their cancer status at the time of diagnosis, and treated age acceleration and other covariates as time-dependent variables using data from Visits 2, 3, and 5. Additionally, we ran a sensitivity analysis and constructed another PAC among two-thirds of all ARIC participants attending Visit 5 (the general population) (“PAC_G”) rather than cancer-free participants. We examined the association between age acceleration for PAC_G and mortality in cancer survivors and cancer-free participants in the remaining one-third participants.

## Results

Among cancer-free participants, Pearson’s r between the new PAC and chronological age was 0.89 (MAE=1.76 years) in the training and 0.75 (MAE=2.19 years) in the test sets (**Figure 2a**). For Lehallier’s PAC, Pearson’s r with chronological age was 0.80 (MAE=2.21) in the training and 0.70 (MAE=2.50) in the test sets (**Figure 2b**). Among cancer survivors, r=0.75 (MAE=2.45) for the new and 0.70 (MAE=2.48) for Lehallier’s PACs (**Figures 2b** and **2d**). Among cancer-free participants (test set) and cancer survivors, the new and Lehalier’s PACs were correlated (r=0.89). According to the KEGG database, proteins included in the new PAC are related to pathways including cytokine-cytokine receptor interaction, metabolic pathways, insulin signaling pathway, and cancer associated pathways (**Supplemental material**).

**Figure 2.**
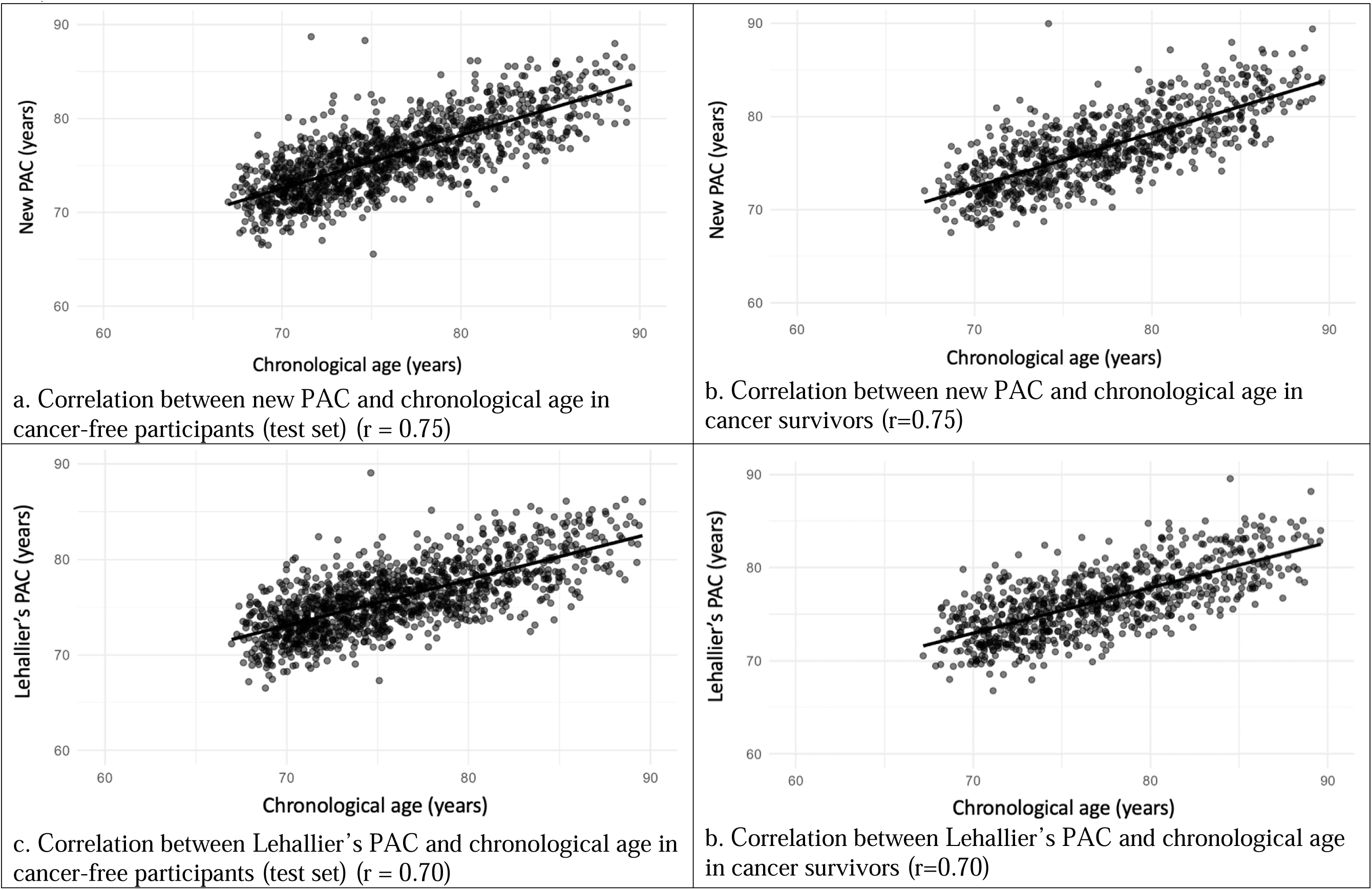
Pearson correlation coefficients (r) of new PAC and Lehallier’s PAC with chronological age in cancer-free participants (test set) and cancer survivors.

Cancer survivors and cancer-free participants (test set) with higher age acceleration for both PACs tended to be White and have prevalent CVD, lower physical activity levels, and lower eGFR (**Table 1**). In addition, cancer-free participants with higher age acceleration for the new PAC were less likely to have diabetes (**Table 1**).

**Table 1.** Visit 5 participant characteristics of across tertiles of age acceleration^a^ in cancer survivors and cancer-free participants (test set)^a^; ARIC.

|  | Cancer survivors |  |  |  |  |  |  |  |
| --- | --- | --- | --- | --- | --- | --- | --- | --- |
|  | New PAC<sup>b</sup> |  |  |  | Lehallier's PAC<sup>c</sup> |  |  |  |
|  | T1 (N=268) | T2 (N=269) | T3 (N=269) | P-value<sup>d</sup> | T1 (N=268) | T2 (N=269) | T3 (N=269) | P-value<sup>d</sup> |
| Range of age acceleration, years | -6.68 to -1.30 | -1.28 to 0.94 | 0.96 to 15.07 |  | -6.81 to -1.30 | -1.29 to 1.02 | 1.03 to 17.02 |  |
| Mean age, years (SD) | 77.06 (4.92) | 76.86 (5.18) | 77.11 (5.39) | 0.84 | 76.97 (5.21) | 76.81 (4.79) | 77.24 (5.48) | 0.63 |
| Male, % | 52.24 | 58.36 | 49.81 | 0.12 | 58.21 | 50.19 | 52.04 | 0.15 |
| White, % | 76.49 | 80.67 | 86.25 | 0.02 | 76.87 | 80.67 | 85.87 | 0.03 |
| Education, % |  |  |  |  |  |  |  |  |
| Less than high school | 12.69 | 12.27 | 18.96 |  | 11.19 | 15.24 | 17.47 |  |
| High school equivalent | 41.42 | 39.40 | 41.26 | 0.10 | 38.81 | 42.75 | 40.52 | 0.15 |
| Greater than high school | 45.89 | 48.33 | 39.78 |  | 50.00 | 42.01 | 42.01 |  |
| Mean BMI, kg/m<sup>2</sup> (SD) | 29.50 (5.26) | 28.92 (5.71) | 27.87 (5.95) | 0.004 | 29.57 (5.19) | 28.95 (5.39) | 27.78 (6.27) | 0.001 |
| Smoking status, % |  |  |  |  |  |  |  |  |
| Current smoker | 4.96 | 6.02 | 7.26 |  | 3.67 | 5.83 | 8.75 |  |
| Former smoker | 54.96 | 58.24 | 54.28 | 0.73 | 56.33 | 56.25 | 55.00 | 0.22 |
| Never smoker | 40.08 | 35.74 | 38.46 |  | 40.00 | 37.92 | 36.25 |  |
| Alcohol intake, % |  |  |  |  |  |  |  |  |
| Current drinker | 52.14 | 55.04 | 53.20 |  | 54.51 | 52.33 | 53.57 |  |
| Former drinker | 29.96 | 29.07 | 28.80 | 0.95 | 30.98 | 28.68 | 28.18 | 0.70 |
| Never drinker | 17.90 | 15.89 | 18.00 |  | 14.51 | 18.99 | 18.25 |  |
| Mean physical activity, score<sup>e</sup> (SD) | 2.63 (0.76) | 2.53 (0.76) | 2.39 (0.81) | 0.003 | 2.65 (0.74) | 2.51 (0.82) | 2.39 (0.77) | 0.001 |
| CVD, % | 21.35 | 29.74 | 34.94 | 0.002 | 16.85 | 29.37 | 39.78 | <0.001 |
| Diabetes, % | 38.81 | 31.23 | 35.69 | 0.18 | 33.21 | 37.92 | 34.57 | 0.50 |
| Aspirin use in the past two weeks, % | 70.79 | 71.91 | 70.15 | 0.90 | 67.42 | 72.01 | 73.41 | 0.28 |
| Mean eGFR, ML/min/1.73 m<sup>2</sup> (SD) | 70.99 (18.13) | 65.19 (18.35) | 59.97 (20.33) | <0.001 | 70.91 (17.89) | 65.64 (18.39) | 59.62 (0.43) | <0.001 |
|  | Cancer-free participants (test set) |  |  |  |  |  |  |  |
|  | New PAC |  |  |  | Lehallier's PAC |  |  |  |
|  | T1 (N=411) | T2 (N=411) | T3 (N=411) | P-value<sup>d</sup> | T1 (N=411) | T2 (N=411) | T3 (N=411) | P-value<sup>d</sup> |
| Range of age acceleration | -9.86 to -1.15 | -1.13 to 1.07 | 1.09 to 15.28 |  | -8.20 to -1.17 | -1.16 to 1.01 | 1.01 to 16.39 |  |
| Mean age, years (SD) | 76.11 (5.15) | 75.47 (5.01) | 76.30 (5.28) | 0.06 | 76.03 (5.14) | 75.80 (5.06) | 76.06 (5.28) | 0.73 |
| Male, % | 43.55 | 40.63 | 40.15 | 0.56 | 39.17 | 43.31 | 41.85 | 0.47 |
| White, % | 75.18 | 80.54 | 82.73 | 0.04 | 75.43 | 79.81 | 83.21 | 0.02 |
| Education, % |  |  |  |  |  |  |  |  |
| Less than high school | 12.90 | 13.63 | 14.67 | 0.89 | 12.40 | 14.88 | 13.90 | 0.44 |
| High school equivalent | 40.87 | 42.82 | 42.05 |  | 43.80 | 38.29 | 43.66 |  |
| Greater than high school | 46.23 | 43.55 | 43.28 |  | 43.80 | 46.83 | 42.44 |  |
| Mean BMI, kg/m<sup>2</sup> (SD) | 29.35 (5.31) | 29.04 (5.77) | 27.62 (5.71) | <0.001 | 29.09 (4.78) | 28.95 (6.11) | 27.99 (5.91) | 0.01 |
| Smoking status, % |  |  |  |  |  |  |  |  |
| Current smoker | 5.78 | 5.54 | 7.22 |  | 5.20 | 6.54 | 6.82 |  |
| Former smoker | 55.10 | 52.91 | 46.95 | 0.25 | 51.51 | 53.95 | 49.43 | 0.64 |
| Never smoker | 39.12 | 41.55 | 45.83 |  | 43.29 | 39.51 | 43.75 |  |
| Alcohol intake, % |  |  |  |  |  |  |  |  |
| Current drinker | 48.46 | 48.06 | 54.09 |  | 46.29 | 51.40 | 52.96 |  |
| Former drinker | 30.77 | 28.68 | 23.75 | 0.21 | 31.46 | 27.73 | 23.92 | 0.18 |
| Never drinker | 20.77 | 23.26 | 22.16 |  | 22.25 | 20.87 | 23.12 |  |
| Mean physical activity, score<sup>e</sup> (SD) | 2.71 (0.81) | 2.65 (0.79) | 2.54 (0.81) | 0.01 | 2.72 (0.79) | 2.62 (0.83) | 2.55 (0.79) | 0.01 |
| CVD<sup>b</sup>, % | 19.95 | 21.65 | 29.76 | 0.002 | 16.55 | 22.93 | 31.87 | <0.001 |
| Diabetes, % | 34.79 | 30.17 | 26.03 | 0.02 | 32.85 | 29.93 | 28.22 | 0.35 |
| Aspirin use in the past two weeks, % | 69.85 | 66.99 | 70.17 | 0.56 | 68.38 | 67.48 | 71.15 | 0.49 |
| Mean eGFR, mL/min/1.73 m<sup>2</sup> (SD) | 73.67 (15.85) | 69.82 (19.27) | 65.53 (18.78) | <0.01 | 72.70 (17.01) | 71.44 (17.47) | 64.90 (19.47) | <0.001 |

Abbreviations: PAC – Proteomic aging clock; SD – standard deviation; CVD – cardiovascular disease; BMI – body mass index; eGFR - estimated glomerular filtration rate.
<sup>a</sup>Age acceleration was calculated as residuals from regressing PAC on chronological age.
<sup>b</sup>The new PAC was developed in cancer-free participants.
<sup>c</sup>Lehallier's PAC was computed using ARIC weights obtained from Ridge regression using proteins available in ARIC.
<sup>d</sup>Physical activity was assessed using a leisure-time sport index that ranged from 1 to 5.
<sup>e</sup>P-values were calculated using chi-square tests for categorical variables and using ANOVA tests for continuous variables.

### Associations between age acceleration and mortality

Among cancer survivors, age acceleration for the new (HR per one SD=1.42, 95% CI=1.24 to 1.62) and Lehallier’s PACs (1.40, 1.22 to 1.61) had similar-sized associations with all-cause mortality in cancer survivors (**Table 2**). These associations were not modified by sex (p-interactions=0.80 for new and 0.73 for Lehallier’s PACs) or race (p-interactions=0.77 for new and 0.47 for Lehallier’s PACs). Age acceleration for the new PAC (HR=1.34, 95% CI=1.09 to 1.64) was associated with cancer mortality in cancer survivors, but not for Lehallier’s PAC (1.19, 0.93 to 1.51). Both PACs were positively but insignificantly associated with CVD mortality in cancer survivors (**Table 2**).

**Table 2.** Associations of age acceleration with all-cause mortality and CVD mortality among cancer survivors and cancer-free participants (test set), and cancer mortality among cancer survivors; ARIC.

|  |  | No. of deaths | Total person-years | HR (95% CI)<sup>a</sup> per 1 SD<sup>b</sup> increase in age acceleration<sup>c</sup> |  |
| --- | --- | --- | --- | --- | --- |
|  |  |  |  | New PAC<sup>d</sup> | Lehallier's PAC<sup>e</sup> |
| All-cause mortality | cancer survivors | 272 | 4,963 | 1.42 (1.24, 1.62) | 1.40 (1.22, 1.61) |
|  | cancer-free participants (test set) | 224 | 8,247 | 1.50 (1.28, 1.76) | 1.61 (1.39, 1.87) |
|  | p-interaction |  |  | 0.62 | 0.20 |
| CVD mortality | cancer survivors | 75 | 4,963 | 1.16 (0.88, 1.52) | 1.15 (0.88, 1.49) |
|  | cancer-free participants (test set) | 82 | 8,247 | 1.42 (1.03, 1.96) | 1.61 (1.22, 2.13) |
|  | p-interaction |  |  | 0.16 | 0.01 |
| Cancer mortality | cancer survivors | 86 | 4,963 | 1.34 (1.09, 1.64) | 1.19 (0.93, 1.51) |

Abbreviations: PAC – proteomic aging clock; CVD – cardiovascular disease; SD – standard deviation; BMI – body mass index; eGFR – estimated glomerular filtration rate; HR – hazard ratio; CI – confidence interval.
<sup>a</sup>Adjusted for chronological age, sex, race- center, education, BMI, smoking status, alcohol intake, physical activity, aspirin use, CVD, diabetes, and eGFR.
<sup>b</sup>SDs for age acceleration were: among cancer survivors=2.59 years and 2.58 years for new PAC and Lehallier's PAC, respectively; and among cancer-free participants (test set)=2.55 years and 2.49 years for new PAC and Lehallier's PAC, respectively.
<sup>c</sup>Age acceleration was calculated as residuals from regressing PAC on chronological age.
<sup>d</sup>The new PAC was developed in cancer-free participants.
<sup>e</sup>Lehallier's PAC was computed using ARIC weights obtained from Ridge regression using proteins available in ARIC.

Age acceleration for the new (HR=1.54, 95% CI=1.05 to 2.25) and Lehallier’s PACs (1.72, 1.13 to 2.64) were associated with all-cause mortality in breast cancer survivors. Additionally, age acceleration for the new PAC was associated with all-cause mortality in colorectal cancer survivors (HR=1.96, 95% CI=1.19 to 3.22), but not for Lehallier’s PAC (1.38, 0.87 to 2.17). Both PACs were positively but insignificantly associated with all-cause mortality in prostate cancer survivors (**Table 3**). In the same group of participants, additional adjustment for stage at diagnosis did not change the direction of these associations (**Table 3**).

**Table 3.** Associations of age acceleration with all-cause mortality among breast, prostate, and colorectal cancer survivors; ARIC (2011-2019)

| **All breast, prostate, and colorectal cancer survivors** |  |  |  |  |  |
| --- | --- | --- | --- | --- | --- |
|  | Multivariable-adjusted model<sup>a</sup> | No. of deaths/No. of survivors | Total person-years | HR (95% CI) per 1 SD<sup>b</sup> in age acceleration<sup>c</sup> |  |
|  |  |  |  | New PAC<sup>d</sup> | Lehallier's PAC<sup>e</sup> |
| All breast cancer survivors | Without adjustment for stage | 46/169 | 1,114 | 1.54 (1.05, 2.25) | 1.72 (1.13, 2.64) |
| All prostate cancer survivors | Without adjustment for stage | 88/255 | 1,551 | 1.19 (0.92, 1.53) | 1.28 (0.97, 1.69) |
| All colorectal cancer survivors | Without adjustment for stage | 35/78 | 412 | 1.96 (1.19, 3.22) | 1.38 (0.87, 2.17) |
| **Breast, prostate, and colorectal cancer with information about stage at diagnosis** |  |  |  |  |  |
|  | Multivariable-adjusted model<sup>a</sup> | No. of deaths/No. of survivors | Total person-years | HR (95% CI) per 1 SD<sup>b</sup> in age acceleration<sup>c</sup> |  |
|  |  |  |  | New PAC<sup>d</sup> | Lehallier's PAC<sup>e</sup> |
| Breast cancer survivors with information about stage at diagnosis | Without adjustment for stage | 31/111 | 743 | 2.60 (1.41, 4.79) | 2.63 (1.43, 4.83) |
|  | Additional adjustment for stage | 31/111 | 743 | 2.68 (1.44, 4.99) | 2.64 (1.43, 4.85) |
| Prostate cancer survivors with information about stage at diagnosis | Without adjustment for stage | 80/228 | 1,400 | 1.19 (0.91, 1.56) | 1.27 (0.95, 1.71) |
|  | Additional adjustment for stage | 80/228 | 1,400 | 1.25 (0.95, 1.65) | 1.32 (0.97, 1.80) |
| Colorectal cancer survivors with information about stage at diagnosis | Without adjustment for stage | 23/52 | 335 | 1.84 (0.95, 3.58) | 5.29 (1.70, 16.46) |
|  | Additional adjustment for stage | 23/52 | 335 | 1.49 (0.72, 3.06) | 3.97 (1.14, 13.86) |

Abbreviations: PAC – proteomic aging clock; CVD – cardiovascular disease; SD – standard deviation; BMI – body mass index; eGFR – estimated glomerular filtration rate; HR – hazard ratio; CI – confidence interval.
<sup>a</sup>Model was adjusted for chronological age, sex, race-center, education, BMI, smoking status, alcohol intake, physical activity, aspirin use, CVD, diabetes, and eGFR.
<sup>b</sup>SDs for age acceleration were: new PAC = 2.60, 2.45, and 2.52 years for breast, prostate, and colorectal cancer survivors and Lehallier's PAC = 2.42, 2.57, and 2.78 years for breast, prostate, and colorectal cancer survivors.
<sup>c</sup>Age acceleration was calculated as residuals from regressing PAC on chronological age.
<sup>d</sup>The new PAC was developed in cancer-free participants.
<sup>e</sup>Lehallier's PAC was computed using ARIC weights obtained from Ridge regression using proteins available in ARIC.

Among cancer-free participants (test set), age acceleration for the new (HR=1.50, 95% CI=1.28 to 1.76) and Lehallier’s PACs (1.61, 1.39 to 1.87) showed similar-sized associations with all-cause mortality (**Table 2**). These associations were not modified by sex (p-interactions=0.32 for new and 0.23 for Lehallier’s PACs) or race (p-interactions=0.82 for new and 0.32 for Lehallier’s PACs). Age acceleration for the new (HR = 1.42, 95% CI = 1.03 to 1.96) and Lehallier’s PACs [HR=1.61, 95% CI=1.22 to 2.13) was associated with CVD mortality in cancer-free participants (**Table 2**).

Cancer status did not modify the associations with all-cause mortality for either the new or Lehallier’s PACs (p-interactions≥0.20) (**Table 2**). In parallel, results from the time-dependent model (exploratory analysis) showed similar HRs (results not shown) for all-cause mortality among those with and without cancer (p-interaction with cancer status=0.05). The association with CVD mortality was modified by cancer status for Lehallier’s PAC (p-interaction=0.01) but not for new PAC (p-interaction=0.16) but sample size was limited (**Table 2**).

### The general population PAC (PAC_G)

Age acceleration for PAC_G was associated with all-cause mortality in cancer survivors (HR=1.56, 95% CI=1.19 to 2.05) and cancer-free participants (1.57, 1.33 to 1.85). Age acceleration for PAC_G was positively but insignificantly associated with CVD mortality in cancer survivors (HR=1.16, 95% CI=0.50 to 2.72) and cancer-free participants (1.30, 0.99 to 1.72) and with cancer mortality in cancer survivors (1.36, 0.30 to 6.21). The directions of the associations for all types of mortality were the same for the new PAC and PAC_G.

## Discussion

In a large prospective cohort of White and Black participants (66-90 years), we developed and validated a new proteomic aging clock (PAC). The correlations between this new PAC and chronological age were the same in cancer-free participants and cancer survivors (r=0.75). Age acceleration for the new PAC showed similar-sized associations with all-cause mortality in cancer survivors and cancer-free participants. Additionally, the new PAC was associated with cancer mortality in cancer survivors, all-cause mortality in breast and colorectal cancer survivors, and CVD mortality in cancer-free participants. We also computed Lehallier’s PAC, which showed the same correlation (r=0.70) with chronological age and similar-sized association with all-cause mortality in cancer-free participants and cancer survivors. Lehallier’s PAC and the new PAC had similar-sized associations with all-cause mortality in both cancer survivors and cancer-free participants and CVD mortality in cancer-free participants, respectively. Moreover, the associations with all types of mortality in cancer survivors and cancer-free participantsfor the PAC developed in the general population (PAC_G) and the new PAC are in the same direction.

In this study, the same correlation with chronological age and similar-sized associations with all-cause mortality in those with and without cancer for both PACs may be explained by the fact that 80% of the cancer survivors in our study have survived at least five years, effectively managing their cancer. Supporting this, a previous ARIC study reported that the overall health of cancer survivors who survived five or more years was comparable to their overall health before their cancer diagnosis.^28^

While the associations with mortality for the new and Lehallier’s PACs were generally similar, a few inconsistencies were observed. For example, we found a significant association between the new PAC and cancer mortality in cancer survivors, but not for Lehallier’s PAC. Furthermore, we found that cancer status modified the association between Lehallier’s PAC and CVD mortality, but not for the new PAC. These variations may be explained by the different aspects of aging captured by these PACs or limited numbers of CVD deaths.

Our study was the first to examine the association between PACs and mortality in cancer survivors. We found that PACs were associated with all-cause mortality in cancer survivors. Since there are no studies tested PACs in cancer survivors, we compared our results to a few studies using epigenetic clocks. PACs’s associations with mortality in our study seems stronger compared to the results from the Melbourne Collaborative Cohort Study (MCCS), where Dugue *et al.* found age acceleration for Hannum clock was associated with all-cause mortality (HR per 5-year=1.06, p=0.01).^7^ However, the MCCS had a longer follow-up time of up to 25 years compared to up to 8 years in ARIC. A longer follow-up may introduce regression dilution bias,^29^ resulting in weaker associations. When restricting follow-up to four years, the association with all-cause mortality for the new PAC (HR=2.02, 95% CI=1.50 to 2.70) was similar to the four-year mortality risk for GrimAge (2.09, 1.45 to 3.00) but stronger than those for Hannum, Horvath, DNAm PhenoAge (HRs=1.27-1.55) in our previous study in the Health and Retirement Study (HRS). In our study, age acceleration for the new and Lehallier’s PACs was associated with all-cause mortality in breast cancer survivors even after additional adjustments for stage at diagnosis. Additionally, age acceleration for the new PAC was associated with all-cause mortality in colorectal cancer survivors in the fully-adjusted model without adjustment for stage (HR=1.96, 95% CI=1.19 to 3.22); this association is stronger than those for epigenetic clocks without adjustment for stage (HRs=1.15-1.30) reported in a previous study.^8^ In summary, PACs showed either similar or stronger associations with mortality in cancer survivors compared to epigenetic clocks.

There is a growing concern about accelerated aging among the rapidly increasing number of cancer survivors. Clinical assessments of aging, such as frailty and comprehensive geriatric assessment, are time-consuming and difficult to collect, particularly in elderly cancer survivors. PACs can be easily measured with a small drop of blood, which may facilitate the measurement of the aging process in cancer survivors in clinics in the future. Although the associations between age acceleration and mortality are similar in those with and without cancer, PACs may be especially important for cancer survivors to estimate their biological age and predict their mortality risk. This information can educate patients about the distinction between biological age and chronological age and inform physicians on the use of lifestyle and therapeutic anti-aging interventions. Targeting biological age is advantageous because it can simultaneously delay the development of multiple age-related diseases.^30^ Additionally, PACs could be applied in clinical trials to test the impact of cancer treatment on the aging process. Lastly, proteins comprising PACs may serve as targets for novel anti-aging drugs for cancer survivors.

The first limitation of our study is that cancer survivors markedly differed by time since cancer diagnosis relative to blood collection time (ranged 2-25 years). However, time since diagnosis did not modify the association between age acceleration and mortality (p-interaction with time since diagnosis=0.36). Second, the numbers of cancer survivors and deaths, especially CVD and cancer deaths, were limited in our study; thus, large-scale studies are needed to validate the results, especially for specific cancer types. Third, detailed information about cancer treatment is lacking, but our goal was to investigate long-term cancer survivors who survived beyond their first course of treatment. Moreover, stage at diagnosis was not available for all cancer survivors. However, additional adjustment for stage did not change the direction of the associations between age acceleration and all-cause mortality in survivors of breast, prostate, or colorectal cancers. Further, the possibility of protein degradation during long-term storage (for 18 years) cannot be excluded. However, the blood samples were frozen right after their collection and have never been thawed, reducing the possibility of degradation.^31^ The strengths of this population-based study include the prospective study design and validated information about the date of cancer diagnosis, cancer site, and stage at diagnosis. Another strength is that we compared the associations with all-cause and CVD mortality in cancer survivors and cancer-free participants.

In conclusion, a new PAC, developed in White and Black individuals, was associated with mortality in cancer survivors and cancer-free participants. The proteins comprising new PAC hold promise as potential targets for anti-aging drugs for cancer survivors. Future studies are needed to confirm our results in a larger population of cancer survivors.

## Data Availability

Data Availability Statement
The ARIC datasets are available through BioLINCC, with appropriate study approvals consistent with NIH policies. Data request forms through BioLINCC can be accessed at https://biolincc.nhlbi.nih.gov/studies/aric/.

## Acknowledgment

The Atherosclerosis Risk in Communities study has been funded in whole or in part with Federal funds from the National Heart, Lung, and Blood Institute; National Institutes of Health; Department of Health and Human Services, under Contract nos. (75N92022D00001, 75N92022D00002, 75N92022D00003, 75N92022D00004, 75N92022D00005). SomaLogic Inc. conducted the SomaScan assays in exchange for the use of ARIC data. This work was supported in part by NIH/NHLBI grant R01 HL134320. Cancer data in ARIC are also supported by the National Cancer Institute (U01 CA164975 and NU58DP007114). Cancer incidence data have been provided by the Maryland Cancer Registry, Center for Cancer Surveillance and Control, Department of Mental Health and Hygiene, 201 W. Preston Street, Room 400, Baltimore, MD 21201. We acknowledge the State of Maryland, the Maryland Cigarette Restitution Fund, and the National Program of Cancer Registries (NPCR) of the Centers for Disease Control and Prevention (CDC) for the funds that helped support the availability of the cancer registry data. The authors thank the staff and participants of the ARIC study for their important contributions. This research was also supported by the National Institutes of Health’s National Center for Advancing Translational Sciences, grant 1UM1TR004405 and the National Cancer Institute, grant R01CA267977. The content of this work is solely the responsibility of the authors and does not necessarily represent the official views of the National Institutes of Health.

