## Supplemental Methods for "Proteomic Aging Clocks and the Risk of Mortality among Longer-Term Cancer Survivors in the Atherosclerosis Risk in Communities (ARIC) Study"

Study population

The ARIC study is a prospective cohort of mostly White and Black females and males.[1,2] At Visit 1 (1987-89), the study recruited 15,792 volunteers aged 45-64 years from four study centers: Washington County, Maryland; the northwest suburbs of Minneapolis, Minnesota; Jackson, Mississippi; and Forsyth County, North Carolina. Participants from Minnesota and Maryland were primarily White, while recruitment in Mississippi was restricted to Black residents. Institutional review boards at each participating center granted approval for the ARIC study, and all study participants provided written informed consent. Over the course of the study, ten visits have been completed.[1] Additionally, ARIC participants have undergone annual telephone follow-ups from 1987 to 2012 and semi-annual telephone follow-ups after 2012. The response rates ranged from 90% to 99% for the annual follow-up calls and 83% to 90% for semi-annual follow-up calls among living participants who have not withdrawn consent to be contacted.[2]

Assessment of cancer cases

Cancer cases were ascertained through 2015 using state cancer registries in the four study centers, complemented by the abstraction of medical records and hospital discharge summaries.[19] An expert panel systematically adjudicated all cases of most common cancers, such as breast, colorectal, lung, and prostate cancers. For adjudicated cases, stage at diagnosis was determined using the pathologic TNM stage (tumor extent, lymph node involvement, presence of metastasis) from the cancer registry or medical records. In instances where pathologic TNM stage was not available, stage at diagnosis was determined according to Surveillance, Epidemiology, and End Results (SEER) summary stage.[19]

Assessment of mortality

Deaths were identified through annual and semi-annual telephone follow-ups with participants or their proxies, state records, and linkage to the National Death Index.[3] The date and cause of death were verified through a review of death certificates. All-cause mortality was defined as death from any cause. Cause-specific deaths due to CVD and cancer were defined based on the underlying cause of death: *International Classification of Diseases*, *Ninth Revision*, codes (ICD-9 codes) 390–459 or *International Classification of Diseases, Tenth Revision*, codes (ICD-10 codes) I00–I99 for CVD deaths. ICD-9 code 140-239 or ICD-10 codes C00-C97 for cancer deaths.

Assessment of participant characteristics

Participants' characteristics of interest included demographic, lifestyle, and medical factors, including chronological age, sex, race, study center, education, cigarette smoking status, alcohol intake, body mass index (BMI), aspirin use, diabetes, CVD, and estimated glomerular filtration rate (eGFR). Participants reported their educational attainment at Visit 1, while all the other characteristics of interest were collected at each visit.[10] During each visit, participants provided information on smoking, alcohol intake, as well as medication use, and underwent a physical examination that included height and weight measurements. eGFR was calculated based on serum creatinine and cystatin C, incorporating age and sex.[11] The calculation for BMI and the definitions for diabetes and CVD have been detailed in our previous studies.[12]

Protein measurement

The protocol for blood sample collection, processing, and storage was specifically designed to minimize the spontaneous biochemical reactions following blood collection (https://aric.cscc.unc.edu/aric9/sites/default/files/public/visitdocuments/v5/Manual%207%20Biospecimen%20Collection%20and%20Processing.pdf) and aligns with the recommended practice for proteomic data analysis in epidemiological studies.[4-6] Briefly, after venipuncture, blood samples were promptly placed in an ice water bath. Centrifugation was conducted within 10 minutes after venipuncture at room temperature (15-25 °C). Following centrifugation, the aliquots were stored at −80 °C within 90 minutes from venipuncture and were never thawed before this analysis.

Plasma samples collected at Visit 2 (1990-92), Visit 3 (1993-95) and Visit 5 (2011-13) were used in this study. Plasma proteins were measured using the SomaScan assay. The details of the SomaScan assay and the data normalization process have been described previously.[6-8] Proteins with a Bland-Altman coefficient of variation (CVBA) greater than 50% or a variance of less than 0.01 on the log scale, or binding to mouse Fc-fusion, contaminants, or non-proteins were excluded.[9] After exclusion, CVBA for split samples was 6% at Visit 2, 12% at Visit 3, and 7% at Visit 5. Protein measures were expressed in relative fluorescent units (RFU) and were log2-transformed to correct for skewness.

### Statistical analysis

#### *Construction of PACs*

The new PAC was developed using cancer-free participants following the methodology described in our previous study.[12-14] In brief, using the training set, we applied elastic net regression (alpha = 0.5 and lambda value was selected based on 10-fold cross-validation) to train PAC against chronological age. A total of 619 aptamers was selected by the elastic net regression, and new PAC was constructed as a weighted sum of proteins: $\beta_0 + \sum_{i=1}^{619} \beta_i \times aptamer_i$, where $aptamer_i$ represents the level of *ith* aptamer. The intercept ($\beta_0$) and weights ($\beta_i$) were estimated using the training set. To compute Lehallier's PAC, using the training set, we applied Ridge regression to estimate the ARIC weights for the available aptamers in ARIC. We chose to estimate ARIC weights instead of using the published weights because ARIC included only 415 aptamers out of those 491 aptamers reported in Lehallier's PAC.[15]

#### *Associations between PACs and mortality*

To capture the effects of PACs that are independent of chronological age, we calculated age acceleration for each PAC as residuals after regressing each PAC on chronological age.[16] We used Cox proportional hazards regression to calculate hazard ratios (HRs) and 95% confidence intervals (CIs) for the associations of age acceleration with all-cause mortality and CVD mortality in cancer survivors and in cancer-free participants (test set), and cancer mortality in the cancer survivors. For the analysis of CVD mortality and cancer mortality, we treated deaths from other causes as competing events following the Fine and Gray method.[17,18] Age acceleration was modeled as a continuous variable as no nonlinear association was observed when applying cubic splines. For each participant, the total person-years were determined from the date of Visit 5 until death, loss to follow-up, or the end of follow up (either December 31, 2017 for participants from Mississippi or December 31, 2019 for participants from other centers), whichever occurred first. The proportional hazards assumption, assessed by modeling an interaction between age acceleration and follow-up time, was not violated in any regression model. We adjusted for chronological age, sex, race-center (a five-category variable: Black participants from Mississippi; Black participants from any other centers; White participants from Maryland; White participants from North Carolina; and White participants from Minnesota), education, BMI, smoking status, alcohol intake, aspirin use, CVD, diabetes and eGFR (fully-adjusted model). In this study, we reported results only for a fully-adjusted model because the HRs for mortality were very similar in demographic-adjusted and fully-adjusted models. We compared the associations in cancer survivors and cancer-free participants by calculating the p-value for interaction between age acceleration and cancer status in the fully-adjusted model.
